# Rapid COVID-19 Diagnosis Using Deep Learning of the Computerized Tomography Scans

**DOI:** 10.1101/2020.12.20.20248582

**Authors:** Hamed Tabrizchi, Amir Mosavi, Akos Szabo-Gali, Laszlo Nadai

## Abstract

Several studies suggest that COVID-19 may be accompanied by symptoms such as a dry cough, muscle aches, sore throat, and mild to moderate respiratory illness. The symptoms of this disease indicate the fact that COVID-19 causes noticeable negative effects on the lungs. Therefore, considering the health status of the lungs using X-rays and CT scans of the chest can significantly help diagnose COVID-19 infection. Due to the fact that most of the methods that have been proposed to COVID-19 diagnose deal with the lengthy testing time and also might give more false positive and false negative results, this paper aims to review and implement artificial intelligence (AI) image-based diagnosis methods in order to detect coronavirus infection with zero or near to zero false positives and false negatives rates. Besides the already existing AI image-based medical diagnosis method for the other well-known disease, this study aims on finding the most accurate COVID-19 detection method among AI methods such as machine learning (ML) and artificial neural network (ANN), ensemble learning (EL) methods.

## I. Introduction

COVID-19 is a global pandemic that collapsed the healthcare systems in most countries. In the year 2020, people all over the world witnessed the news of the death of their fellow human beings from many world news agencies. Furthermore, this pandemic event has affected the operations of healthcare facilities. The medical centres witnessed increases in patients who are needing care for a respiratory illness that could be COVID-19 (+) or COVID-19 (-). The World Health Organization (WHO) advises that all countries to consider the importance of the test because the isolation of all confirmed cases and also mild cases in health centers is able to prevent transmission and provide acceptable care. One of the pivotal reasons for the need to use intelligent systems in the process of diagnosing this disease (taste) is the easy transmission of this disease among people in a community or even health facilities [1,2]. Since most of the excited test needs a lot of time to generate the result compared to the time for spreading virus among people, chest X-Ray or Computer Tomography (CT) scan images of COVID19 is used to provide a rapid and efficient way to test the COVID-19 suspected individuals. It is an undeniable fact that artificial intelligence plays a central role in making human daily life more convenient than the past. The advantage of AI methods is their ability to interpret and understand the digital images in order to identify and classify objects. For this reason, many researchers in the world of artificial intelligence have drawn attention to research on the data obtained from patients who infected with COVID-19. Sachin Sharma [3] presents a study that aims to discuss the importance of machine learning methods to distinguish COVID-19 infected regarding their lung CT scan images. Nripendra Narayan Das et al. [4] use chest X-rays in order to find some radiological signatures of COVID-19 by using deep learning of the chest CT scans. Aayush Jaiswal et al. [5] use the pre-trained deep learning architectures (DenseNet201) along with deep transfer learning in order to provide an automated tool that aims to detect COVID-19 positive and negative infected patients based on chest CT images. Xueyan Mei et al. [6] combine chest CT records including the patients’ essential symptoms. In this pioneer research the interaction between the chest CT and the clinical symptoms is conducted through basic machine learning methods, i.e., SVM, random forest, MLP, and deep learning to accurately predict COVID-19. In an alternative approach, Pinter et al. [7] present the hybrid machine learning method of ANFIS and MLP to predict mortality rate of COVID-19 patients. Sina F. Ardabili et al. [8] review a wide range of machine learning models to forecast the COVID-19 outbreak. Their study presents a number of suggestions to demonstrate the potential of machine learning for future research.

In a nutshell, the main motivation of this paper is to find the most accurate intelligent approach for detecting COVID-19. In other words, we use state-of-the-art learning models in order to classify positive and negative COVID-19 suspected individuals with regard to their captured chest X-Ray or CT scan images.

The rest of this paper is outlined as follows. Section 2 reviews Machine leaning-based models. Section 3 compares the performances of the described and implemented machine learning models. Finally, Section 4 draws conclusions and offers some suggestions for the end-users of medical intelligent systems.

## II. Brief review of Machine leaning-based models

AI has the potential to improve medical imaging capabilities and patient diagnosis. Using ML, ANN, and ensemble learning methods for medical image recognition is a core component of computer vision in this widespread study area. ML methodology works based on the cognitive learning methods to advance an intelligent code without use of conventional programing techniques. The performance of ML algorithms’ is associated with other mathematical techniques and improved by experience [4]. Generally, ML uses historical data to make decisions and uncover hidden insights [5]. In image-based diagnosis problems, the ML models are advanced to be able to learn from medical records. This process is often done through developing insight into the patterns within complex imaging [6]. The following subsection describes the basic and ensemble AI-based image classifier methods in a brief way.

### A. Single models

In the following, the basic classifiers employed for diagnosing COVID-19 are introduced.

- *Support Vector Machine (SVM)* is one of the commonly-used algorithms in research and industry, taking its power from machine learning algorithms. The main advantage of this algorithm is its ability to deal with non-linear problems. SVM can be used to solve nonlinear classification problems by transforming the problem using the kernel method which makes SVM calculation in the higher dimension. Vapnik was first introduced SVM in 1995[]. He used the Statistic Learning Theory (SLT) and Structural Risk Minimization (SRM) to introduce this concept. SVM can be effectively employed in classification, regression, and nonlinear function approximation problems [9,10].
- *Naive Bayes (NB)* is a well-known probabilistic classification algorithm that applies Bayes’ Theorem with an assumption of strong (naive) independence among predictors (a set of supervised learning algorithms). During the process of constructing classifiers (training), the NB model needs a small amount of training data to estimate the vital parameters [11]. In other words, the previous probability of each class is estimated by calculating the conditional probability density function and the posterior probability. Eventually, the final prediction is made for the class that has the largest posterior probability.
- *Artificial neural networks (ANN)* are computing systems that widely used for image-based medical diagnosis problems. In fact, ANN draws inspiration from biological neural systems and creates an interconnected network of ‘neurons that process information. These models consist of several processing elements that reproduce input data in a hierarchical structure. During the training process, a corresponding weight (for each input data) must be iteratively estimated and adjusted. Due to the variation of connections between layers in an ANN, the architecture of networks is able to design variously. Deciding the number of layers and nodes in each layer depends on the problem and the amount of training data. For this reason, ANN is a great (flexible) option to deal with different classification, regression, and clustering problems [12,13].
- *Multilayer perceptron (MLP)* is a well-known ANN in which neurons are distributed in thoroughly connected layers. These layers are divided into three groups: input layers, output layers, and hidden layers. The weighted inputs are linearly combined by their corresponding neuron; then, the results are transferred through a nonlinear activation function. Usually, a gradient-descent algorithm called back-propagation is used to train an MLP. In this algorithm, a maximum error is defined to be used as a criterion to stop the iterative weight update process [14].
- *CNN* is a deep neural network that is commonly applied to process large scale images. As same as the other ANN, CNN is a network that includes several layers. In CNN represents a sequential connection between the layers. As the output of the previous layer is interconnected with the input of other layer. However, unlike the other fully connected neural network, in this network, the neurons in one layer do not connect to all the neurons in the next layer. The main powerful part of CNN is the convolution layer [15,16].

### B. Ensemble models

Ensemble models are able to scale up the performance of classification and regression processes. Boosting and Bagging are the most widely-used ensemble learning frameworks in science literature. Bagging ensembles create subsets and ensemble estimates using Bootstrap re-sampling and a mean combiner respectively. Boosting ensemble models train a number of individual models in a sequential way. A way that provides an opportunity for each model to learns from mistakes made by the previous model [17].

- *AdaBoost (Adaptive Boosting)* is an ensemble learning algorithm that can be used in conjunction with many other types of learning algorithms to improve performance. AdaBoost initially created to enhance the performance of binary classifiers. The main idea of AdaBoost is about using an iterative approach in order to learn from the mistakes of weak classifiers, and turn them into strong ones. In fact, AdaBoost learns from the mistakes by increasing the weight of misclassified data points [18].
- *Gradient boosting decision tree (GBDT)* is an ML algorithm, which produces a prediction model in the form of an ensemble of weak prediction models (decision trees). Gradient Boosting learns from the residual error (directly), rather than update the weights of data points [19].

In a nutshell, Table I indicates the advantages and disadvantages of all mentioned machine leaning models.

**TABLE I.** ADVANTAGES AND DISADVANTAGES OF ML MODELS

| Models |  | Advantages | Disadvantages |
| --- | --- | --- | --- |
| Single models | SVM | Effective performance in high dimensional spaces | Doesn't perform well with large data set (required high training time) |
|  | NB | Easy and fast class prediction in test datasets | Need to calculate the prior probability |
|  | MLP | Adaptive learning, coefficients can easily be adapted | It requires much more training data than traditional machine learning algorithms |
|  | CNN | Automatically detects the important features (Feature Extraction), uses convolution of image and filters to generate invariant features | Unexplained functioning of the network (Black box), require processors with parallel processing power |
| Ensemble models | AdaBoost | Less susceptible to the overfitting problem than most learning algorithms | Using too weak classifiers can lead to low margins and overfitting |
|  | GBDT | Lots of flexibility (can optimize on different loss functions) | Too much improvement to minimize all errors can overemphasize outliers and cause overfitting |

## III. Experimental Results

In this section, all described models in the previous section were evaluated and examined based on the datasets that include image data on 980 patients suspected with COVID-19 infection.

The implementation is facilitated under Python using Scikit-Learn and Keras libraries. The experimental results are provided and analyzed in detail by using a standard CPU with the information of Intel Core i5-2.20 GHz with 16 GB RAM.

### A. Data description

This paper evaluates all described models mentioned in the previous section based on two datasets. The first data set includes image data on 430 patients infected with COVID-19. Also, 550 healthy (normal) individuals were randomly selected from the second data set [20]. The following Figure illustrates sample images of both classes.

**Fig. 1.**
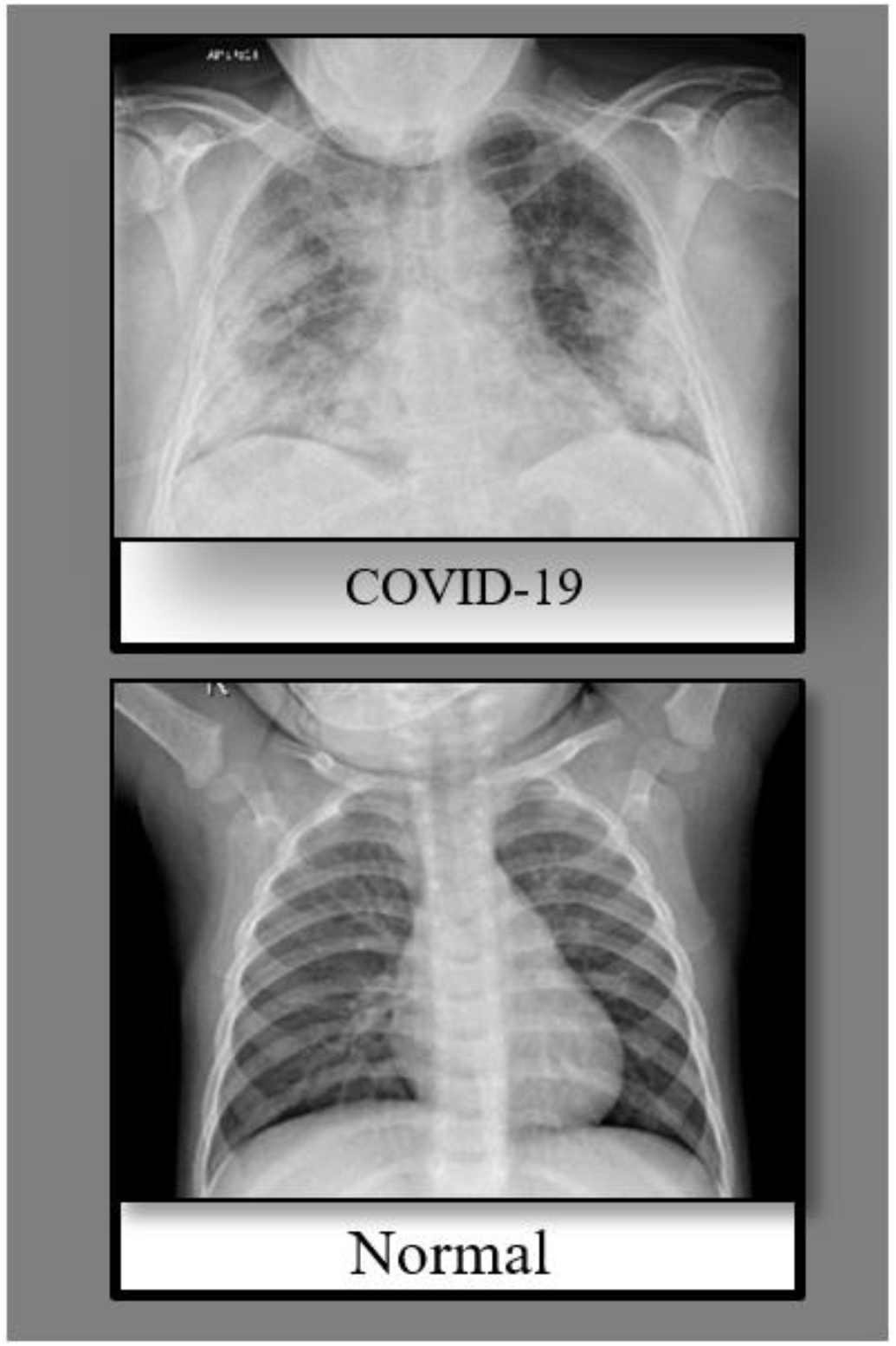
X-ray images of normal and COVID-19 caused patient

### B. Performance evaluation

In the presented study, we split the data images into a training and testing image set. We use 75% of the data as the training data for training the model, the next 25% remaining data were used as testing data. Moreover, all considered models were evaluated by taking advantage of the well-known performance criteria and Matthews correlation coefficient (MCC) [21].

According to the confusion matrix, the formulas for these measurements are described briefly as follows.

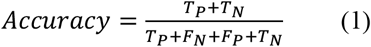

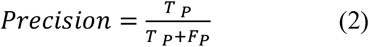

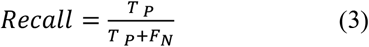

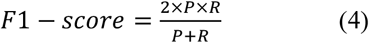

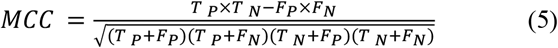

where *F*_*N*_ and *F*_*p*_ present the quantity of the incorrect predictions respectively. *T*_*p*_ and *T*_*N*_ indicate the quantity of correct predictions.

The results of the implemented machine learning algorithms evaluated using the datasets described earlier.

Table. II presents the performance of trained models with regard to the mentioned standard performance criteria.

**TABLE II.**
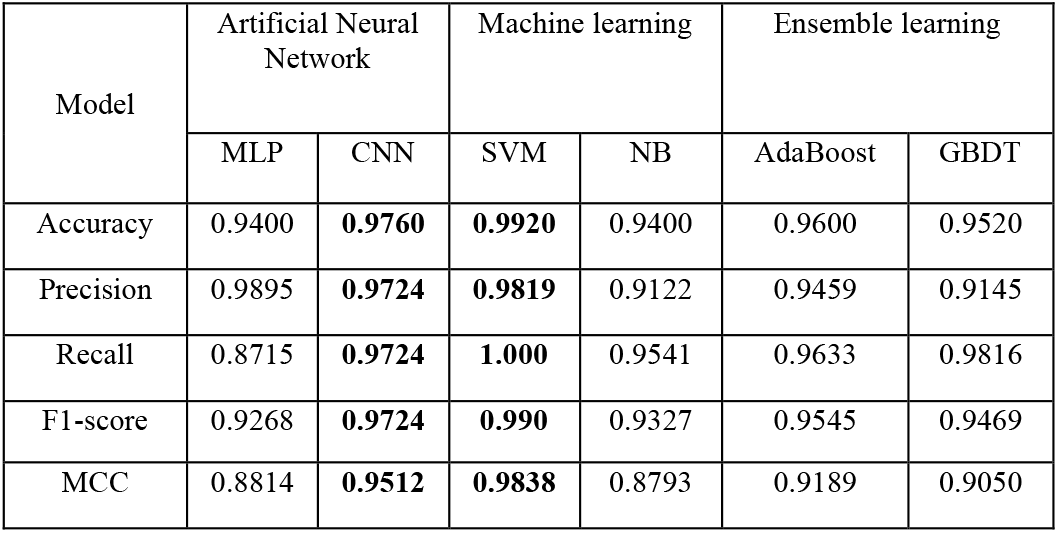
PERFORMANCE COMPARISON BETWEEN COVID-19 DETECTION MODELS

In our experiment, each test set is executed with six different machine learning algorithms. By using the values in the confusion matrix, 5 different statistics (in Equations. (1– 5)) are calculated to measure the efficiency of the algorithms. In this study, the architecture of MLP designed as five fully connected layers with 350, 250,150,50 and 2 number of neurons in each layer, respectively. Except for the last layer, the rectified linear unit (Relu) used in considered MLP network and the last layer use “softmax” activation, which means it will return an array of 2 probability scores (Positive or Negative). In addition, the architecture of CNN designed as four convolution layers with a convolution kernel size of 7×7 to extract the features and each them uses 3×3 average pooling layer or max pooling to prevent the features. The number of convolution in each convolution layer is at least 64. After the max pooling layer, two 512-dimensional fully connected layer are added, along with dropout layer in order to prevent overfitting problem. In Table II, the results show that SVM outperforms the other models. The Table includes the performance of CNN which has provided a 97% accuracy rate. In addition, it is reported an 99% accuracy for the SVM model. Sine image-based diagnosis SVM model has to deal with nonlinear pattern of data, we considered RBF kernel for SVM. Furthermore, the confusion matrix for the considered learning models is constructed in Table III.

**Table. III.** CONFUSION MATRIX

| Model | Confusion matrix |  | Predicted |  |
| --- | --- | --- | --- | --- |
|  |  |  | P | N |
| MLP | Actual | P | 140 | 1 |
|  |  | N | 14 | 95 |
| CNN | Actual | P | **138** | 3 |
|  |  | N | 3 | **106** |
| SVM | Actual | P | **139** | 2 |
|  |  | N | 0 | **109** |
| NB | Actual | P | 131 | 10 |
|  |  | N | 5 | 104 |
| AdaBoost | Actual | P | 131 | 10 |
|  |  | N | 2 | 107 |
| GBDT | Actual | P | 135 | 6 |
|  |  | N | 4 | 105 |

## IV. Conclusions

Rapid diagnosis of COVID-19 symptoms is of utmost importance. This paper implemented COVID-19 detection models by using six different machine learning algorithms, as SVM, NB, GBDT, AdaBoost, CNN and MLP based on the datasets that include image data on 980 patients suspected with COVID-19 infection. The main purpose of this study was to introducing image-based ML methods for developers and end-users of intelligent medical systems in a comprehensive way. We compared the performance of the state-of-the-art models to show that how important is the ML image-based models reliability to diagnose diseases. The experimental results and discussions proved that the SVM with RBF kernel outperforms other existing methods.

**TABLE IV.** GLOSSARY OF ACRONYMS

|  |  |
| --- | --- |
| Artificial intelligence | AI |
| Artificial neural networks | ANN |
| Computer Tomography | CT |
| Convolutional neural networks | CNN |
| Ensemble learning | EL |
| Gradient boosting decision tree | GBDT |
| Machine learning | ML |
| Matthews correlation coefficient | MCC |
| Multilayer perceptron | MLP |
| Naive Bayes | NB |
| Statistic Learning Theory | SLT |
| Structural Risk Minimization | SRM |
| Support Vector Machine | SVM |
| World Health Organization | WHO |

## Data Availability

Data is available upon request.

## Acknowledgment

We acknowledge the financial support of this work by the Hungarian State and the European Union under the EFOP-3.6.1-16-2016-00010 project and the 2017-1.3.1-VKE-2017-00025 project. The research presented in this paper was carried out as part of the EFOP-3.6.2-16-2017-00016 project in the framework of the New Szechenyi Plan. The completion of this project is funded by the European Union and co-financed by the European Social Fund. We acknowledge the financial support of this work by the Hungarian State and the European Union under the EFOP-3.6.1-16-2016-00010 project. We acknowledge the financial support of this work by the Hungarian-Mexican bilateral Scientific and Technological (2019-2.1.11-TÉT-2019-00007) project. The support of the Alexander von Humboldt Foundation is acknowledged.

